# Implementing the MATILDA intervention to enhance community participation for older adults with intellectual disability: a feasibility randomised controlled trial and RE-AIM guided process evaluation

**DOI:** 10.64898/2026.01.16.26344257

**Authors:** L Taggart, A Campbell, I Doherty, K Ensue, H Ahmed, A Hanna, L Armstrong, J Ding, B Bunting, A Ryan, C O’Neill, M Clarke, B Leahy, S Willis, J Schofield, R Stancliffe, A Hassiotis

## Abstract

**Aim:** The aim of this study was to: 1) assess the practicality of conducting a feasibility randomised controlled trial and 2) undertake a process evaluation of MATILDA guided by the RE-AIM Framework (Acceptability, Reach, Effectiveness, Adoption, Implementation and Maintenance).

**Methods:** This was a 2-arm, single-blind, randomised feasibility trial with 1:1 allocation conducted in the UK. The intervention lasted 6-months. Clinical outcome variables (quality of life, anxiety, loneliness) were collated at baseline, 6- and 12-months post intervention. The RE-AIM framework was used to undertake the process evaluation. A health economic evaluation was also explored.

**Results:** We recruited 57 participants of our target population of 64 (89%), 28 participants were randomly allocated to MATILDA and 29 to an active control group. We lost three participants in the intervention arm prior to commencement of the intervention. We were able to match all 25 participants to a community group, most with a mentor(s). With regards reach and implementation, most older adults with an intellectual disability were still attending their community groups at 6-months (N= 18, 72%) and at 12-months (N= 17, 94%). There was a slight decrease in the anxiety scores for the participants in Matilda over the 6- and 12-months follow-up periods compared to the control group. MATILDA was found to be clearly acceptable and adoptable by the community groups and mentors. There were some challenges identifying and recruiting community groups and getting mentors to complete their questionnaires.

**Discussion:** MATILDA appears to be a promising community-based intervention to support older adults with an intellectual disability that promotes social inclusion, quality of life, and wellbeing. This study advances our empirical knowledge in understanding the contextual factors, mechanisms and outcomes in developing and delivering a community intervention for this population.

Trial feasibility and process evaluation of the MATILDA intervention to support older adults with an intellectual disability to integrate into local community groups using the RE-AIM Framework

## Background

Older adults commonly experience increased physical and mental health needs alongside heightened risks of social isolation and loneliness (1–4). Loneliness in later life is strongly associated with poorer wellbeing, depression and reduced quality of life (5–7), leading to increased health and social care utilisation (8,9). Facilitating participation in local community groups is known to promote social connectedness, reduce loneliness and improve wellbeing (5,10).

One model for enhancing social connection is the use of community-based interventions such as befriending schemes, where volunteers support individuals with limited social networks to engage socially and participate in community life, thereby promote a sense of belonging to their communities. Evidence in the general population suggests befriending may reduce social isolation, loneliness and depression (11), benefit volunteers through enhanced empathy and social awareness (12), and strengthen community cohesion while potentially reducing public sector costs (13). Another model was community-based intervention groups focusing on arts / activities / hobbies, social engagement and peer support thereby promoting a sense of belonging (14). Further, these community-based groups have been found to provide sustained social interactions which were core to maintaining long-term well-being (15).

### Older adults with an intellectual disability

People with an intellectual disability are living longer, although they experience accelerated and earlier ageing, often beginning from approximately 45 years of age, and earlier still (at 40 years) among people with Down syndrome (16–18). Older adults with an intellectual disability frequently experience social exclusion, reduced access to mainstream social opportunities, and fewer friendships outside disability services (19,20). They also report disproportionately high levels of loneliness, depression, and low quality of life (3, 4, 9,21,22), despite consistently expressing a desire to build friendships and participate in their communities (19, 20).

A systematic review reported loneliness among people with intellectual disability at levels of nearly five times the prevalence seen in the general population (22).

Barriers included communication challenges, limited opportunities to develop social networks, and societal attitudes that reinforce exclusion (23).

Although befriending schemes exist for people with intellectual disability, evidence of their effectiveness remains limited. A small feasibility study suggested potential benefits but lacked statistical power (24), while a feasibility randomised trial in adults with intellectual disability in London, UK struggled to recruit sufficient participants and did not progress to a full trial (25). Furthermore, befriending schemes may not promote sustainable social participation: once a volunteer withdraws, social connections may not continue. Thus, there is no robust evidence that befriending schemes improves clinical or cost outcomes for people with an intellectual disability, and existing befriending schemes may not build enduring community integration. Mnay people with intellectual disability attend community-based groups that are offered only to other people with an intellectual disability. However, there is now growing research that is examining how people with intellectual disability engage in non-disability community-based groups.

Policy guidance emphasises inclusion of people with intellectual disability in their local community groups (26–29). UK guidelines from the National Institute for Health and Care Excellence (NICE) (NG96) recommended that older people with an intellectual disability should have equal access to local community-based groups (18). However, there is limited evidence on how best to operationalise these recommendations.

### The MATILDA Intervention

MATILDA (Managing Activities Together to Involve older adults with a Learning Disability in their Local Areas) is a novel community-based, mentor-supported intervention that promotes social inclusion, quality of life and wellbeing. MATILDA is based on the Australian “Transition to Retirement” intervention, which enabled older adults with an intellectual disability to join community-based groups for six-months with volunteer support. MATILDA adapts and extends this approach within UK health and social care contexts (30, 31).

This study was guided by the RE-AIM framework (Reach, Effectiveness, Adoption, Implementation, Maintenance) to structure the evaluation of the feasibility, acceptability, and potential sustainability of the MATILDA intervention. RE-AIM provides a pragmatic implementation lens suited to community-based public health interventions and helps identify whether MATILDA could be scaled in routine practice. This study also tested the practicality and acceptability of conducting a definitive full-scale trial to evaluate MATILDA’s effect on social connectedness, well-being, and quality of life.

### Aim

The aim of this study was to: 1) assess the practicality of conducting a feasibility randomised controlled trial (RCT) and 2) undertake a process evaluation of MATILDA guided by the RE-AIM Framework (Acceptability, Reach, Effectiveness, Adoption, Implementation and Maintenance) (32).

### Objectives

1. Determine feasibility outcomes (eligibility, recruitment, consent, randomisation, matching to community groups and mentors, attendance, retention, dropout)
2. Explore stakeholder views (older adults with intellectual disability, mentors, community groups) on the acceptability of the MATILDA intervention
3. Examine whether mentoring relationships and community participation can be sustained over the study period?
4. Assess appropriateness and acceptability of the clinical outcome measures
5. Explore potential cost-effectiveness.

## Methodology

### Study design and setting

This was a 2-arm, single-blind, randomised feasibility study with 1:1 allocation, which was conducted in two regions of the United Kingdom: one Health and Social Care Trust in Northern Ireland and one Trust in London, England. We approached 95 potential participants and 57 older adults with an intellectual disability agreed to join the study. We randomly allocated them to either the MATILDA intervention plus usual care or usual care with three group recreational activities with other older adults with an intellectual disability (active control arm). The intervention lasted six-months. The primary outcome measures were feasibility outcomes (eligibility, recruitment, consent, randomisation, matching to community groups and mentors, attendance, retention, dropout). Secondary outcome measures focused on the clinical outcomes (i.e. quality of life, anxiety, loneliness). Both primary and secondary outcome measures were collated at baseline, 6- and 12-months post intervention. A process evaluation was undertaken using the RE-AIM framework (32) and a health economic evaluation was also explored. This study was aligned with the CONSORT 2010 extension for pilot and feasibility trials (33).

### Participants and Recruitment

The recruitment process ran from July 2023 to March 2024. There were some delays in the London site, so identification and recruitment of participants commenced in October 2023. Statutory and voluntary disability community groups helped to identify potential adults with an intellectual disability who met the inclusion criteria using a user-friendly information sheet. A member of the research team at each site then met with the potential participant, and if needed a family / paid carer for additional support, to ensure that each participant had the capacity to consent and understood what was involved in the trial, including completing of questionnaires at three time points, randomisation, and if selected participating in a community group for six-months.

### Inclusion Criteria

The inclusion criteria were that each participant had to be at least 45 years of age, living in the community, have a mild / moderate intellectual disability, sufficient communication to engage in a community group, and sufficient mobility to attend the community group. All participants had to have the capacity to give their own written consent.

### Sample Size Calculation

As this was a feasibility trial and the purpose was to provide estimates of key parameters for a larger definitive trial rather than to power this study to detect statistically significant differences, a formal power calculation was not conducted (34). For pragmatic reasons we choose a sample size of 64 potential participants with intellectual disability across two sites: one urban and one rural. This would allow us to adequately test the identification and recruitment processes, ease of use of questionnaires, randomisation, matching to mentor(s) and community groups, intervention processes, and the barriers and enablers to implementing the MATILDA intervention in real world settings.

### Randomisation

Following informed consent, the participants were randomised using a 1:1 ratio to the intervention arm (MATILDA) or an active control arm, stratified by site.

Randomisation was generated by a member of staff based at Ulster University, Northern Ireland not connected to the study: this person was blinded. At the time of randomisation, each participant was allocated a unique Participant Study Number, which was used throughout the study for participant identification. The two research staff conducting the pre and post assessments at each site were not blinded to each participant’s allocated group. It was not possible to blind the participants and mentors to arm allocation.

### MATILDA Intervention

Older adults with intellectual disability were matched to one or two mentors from a local community group. This matching process was managed by a Volunteer Co-Ordinator (VC) in Northern Ireland and London. The VC at each site had to assess each person’s capabilities, motivations, interests / hobbies, financial situation, health co-morbidities and living context (i.e. living on their own, with family or in supported living / residential care) transport arrangements and match the person to a local community-based group within the same locality, with similar interests, and pair them to one or two mentors. The VC supported each adult with an intellectual disability to meet their mentor(s) prior to commencing the group. Then the VC physical accompanied each adult with an intellectual disability to attend their local community group and meeting with their mentor(s), then gradually withdrawing their support over the following weeks. The total length of the MATILDA intervention was six-months.

One of the core responsibilities of the VC was to develop a rapport and trusting connection between the adult with the intellectual disability, their family or/and paid carers, and the community group and mentor(s) to allow the participant to move outside of their safe and protected world of disability services. Participants randomised to MATILDA also received treatment as usual (TAU), ensuring that they did not lose any treatments or care that was standard. All mentors completed a short 2-3 hour training session on the role of a mentor to ensure they were aware of the study purpose and how they as mentors would be able to provide support.

### Active Control Arm

Those in the active control group received TAU and were offered three group recreational activities with other older adults with an intellectual disability within this arm: this included going for a meal, going to the museum, zoo, or cinema. TAU was established at the start of the study and again at the end of the study. The participants in control group completed the same data gathering instruments as the intervention group at baseline, 6- and 12-months.

### Feasibility Outcomes / Process Evaluation

A process evaluation was conducted in line with the Medical Research Council’s Guidelines (35) to assess whether MATILDA was delivered and received as intended, and to understand how trial processes interacted with the implementation context. This included an examination of both feasibility outcomes and clinical outcomes, including the acceptability of outcome measures and the perceived benefits and challenges of implementing MATILDA.

We used the RE-AIM framework (Reach, Effectiveness, Adoption, Implementation, and Maintenance) (32) as part of the process evaluation. Reach refers to the number of older adults with an intellectual disability who engaged with their local community groups and mentor(s) at 6-months with support from the VC. We further explored whether those adults still engaged with their local community groups at 12-months.

Effectiveness assessed the impact of MATILDA on the participants’ quality of life, anxiety, loneliness and community connectedness from baseline to the two follow-up time points using the standardised measures, focus groups and VC reports. The effectiveness data were analysed using descriptive statistics. Adoption explored how the mentors, other community group members and the staff in the disability group homes practically and emotionally supported the adults with disability to engage in their groups. Implementation concerned whether MATILDA was delivered as intended. We collated the attendance records of the adults with intellectual disability at the groups, additional activities undertaken from the mentors and the VC’s attendance logs and fidelity checks, respectively. Maintenance assessed the extent to which the MATILDA’s effects were sustained over time at both the individual level and the organisational level.

By considering these five domains, RE-AIM provided a structured approach to undertaking the process evaluation evaluating both the internal validity and external applicability and supporting generalisability and implementation of the MATILDA intervention into routine practice.

### Clinical outcome measures for the adults with intellectual disability

The secondary outcome measures focused on the clinical outcomes and were collated at baseline, 6- and 12-months post intervention for both arms. We used the WHO QOL-Dis as a measure of health-related quality of life developed by the WHO (36). This 26-item short version consists of two benchmark items on general health (not used in the scoring) (one general QOL item, one general health item) and 24 specific items which generate a total score and four domains: physical health (7 items), psychological health (6 items), social relationships (3 items), and environment QOL (8 items). Each item is answered on a 5-point Likert scale, which assesses the intensity, capacity, frequency, and evaluation of QOL facets with respect to the last two-weeks. The WHOQOL-Dis has been validated in other intellectual disability population studies and found to have good psychometric properties (36). Higher scores mean better QOL.

We used the Glasgow Anxiety Scale for (GAS-ID) which was developed for adults with intellectual disability (37). This is a 27-item self-report scale. It has been used in several studies with adults with disability and found to have good psychometric properties (37). Scores ranged from 0 to 54: score of 13 or over indicates anxiety.

We also used the Modified Worker Loneliness Scale, a 12-item questionnaire that measured Aloneness and Social Dissatisfaction in adults with an intellectual disability (38). It has been used in several other disability studies and found to have good psychometric properties (38). We also used the WHO Wellbeing 5, a short 5-item scale to measure subjective wellbeing (39).

We explored a health economic evaluation to examine issues likely to be encountered in the conduct of a full economic appraisal including sources of data, how best to collect these data and the inclusion of spill-over effects into the analysis. We used the Client Receipt Services Inventory (40) to collate service use and costs for people with an intellectual disability over the 12-months of the study. It was completed by the adults with an intellectual disability and their family / paid carer. We also used the EQ-5D-Y (41) a measure of health-related QOL which provides a description of health using five dimensions (mobility, self-care, usual activities, pain/discomfort, and anxiety/depression). Each domain has three response levels of severity: no problems, some problems, extreme problems. Scores range from 5 (no problems) to 25 (severe problems). This scale has been used previously in studies with adults with disability (42).

We collected data from the mentors at baseline and 6- and 12-months post intervention measuring psychological well-being (Warwick Edinburgh Mental Wellbeing Scale (WEMWBS)) (43), and QOL (WHO QOL DIS) (36) and Attitudes Towards People with a Learning Disability (ATPLD) (44).

### Intervention Schedule

We undertook a series of 1-1 interviews and focus groups with twelve adults with intellectual disability, nine mentors, three community group managers and five paid disability staff fronm residential facilities across the two sites: all were involved in the MATILDA intervention. The interview schedules and data collection were informed by RE-AIM framework (32).

## Data Analysis

Given that this was a feasibility study we undertook no statistical tests as the primary aim was to assess the feasibility of conducting a definitive randomised trial of MATILDA, rather than the effectiveness or efficacy of the intervention (33, 45). With regards health economics, we assessed the feasibility of calculating quality adjusted life years (QALYs) using both the EQ5DY (41) and WHOQOL-DIS (36) and including spill-over effects experienced by mentors and carers in the evaluation (40) using the Client Receipt Services Inventory. The focus groups were audio recorded and transcribed and analysed using RE-AIM (32).

## Ethics

Ethical approval was obtained from the Office of Research Ethics N Ireland (ORECNI) (HSC REC B (Ref 22/NI/067)).

## Results

We approached 95 adults with a mild / moderate intellectual disability and obtained written consent from 57 participants (60%): our target sample was 64 (see Table 1). Reasons for exclusion included: not meeting the inclusion criteria and most participants who did not consent gave reasons such as ‘not interested’ and ‘too busy’. Of the 57 participants who agreed to join the study: 29 were males (50.8%) and 28 were females (49.2%). All were in the age range 45 - 81 years (mean age 56.7 years). 33 participants were recruited from Northern Ireland, and 24 participants were recruited from the London site.

**Table 1:** Characteristics of the sample.

| Baseline Characteristics | Treatment Group |  | Total |
| --- | --- | --- | --- |
|  | MATILDA | Control |  |
|  | n= 28 | n= 29 |  |
| Male | 13 (44.8%) | 16(55.2%) | 29 (50.8%) |
| Female | 15 (53.6%) | 13 (46.4%) | 28 (49.2%) |
| Age (years) | 56.6 (7.1%) | 56.7 (10) | 56.68 (8.6) |
| Ethnicity: White | 26 (92.9%) | 23 (79.3%) | 49 (86%) |
| Ethnicity: Black | 1 (3.6%) | 3 (10.3%) | 4 (7.1%) |
| Ethnicity: Asian | 1 (3.6%) | 2 (6.9%) | 3 (5.2%) |
| Ethnicity: Other | 0 | 1 (3.4%) | 1 (1.7%) |
| Living status: Supported living | 13 (46.4%) | 17 (58.6%) | 30 (52.6%) |
| Living status: Own home | 9 (32.1%) | 6 (20.7%) | 16 (28.1%) |
| Living status: Shared care | 3 (10.7%) | 0 | 3 (5.2%) |
| Living status: With Family | 2 (7.1%) | 1 (3.4%) | 3 (5.2%) |
| Living status: Group home | 1 (3.6%) | 5 (17.2%) | 6 (10.5%) |

mean ( ± SD) for continuous data and the number (% in group) for categorical data

### Baseline characteristics

Table 1 provides a summary of the characteristics of the sample including the participants’ gender, age, ethnicity and living status split by both intervention and control arms. There were no differences between the adults with intellectual disability recruited from either the Northern Ireland or London sites.

We also collected data on the participants’ health: 26.3% (n=15) self-reported they had depression, 24.6% (n= 14) anxiety, 19.3% (n= 11) asthma, 17.5% (n= 10) epilepsy, 12.3% (n= 7) diabetes, 12.3% (n= 7) high blood pressure, 5.3% (n= 3) vision problems, 3.5% (n= 3) had had a stroke and two participants had autism (3.5%).

### Recruitment and retention

Staff within community statutory and voluntary disability services advertised the study on our behalf and spoke to potential participants who met the inclusion criteria. For those participants who verbally expressed an interest, their contact details were passed to the research team who then met the person and/or their family / paid carer. Capacity to consent was assessed and checks were made by the research team to ensure that each participant met the inclusion criteria.

Recruitment took place from July to October 2023 in Northern Ireland, and October 2023 to February 2024 in London (see Table 2). This was an average of approximately 8 participants per month. We recruited 57 participants of our target population of 64 (95%). Because of delays in starting in the London site, recruitment for mentors started in November 2023 and was completed in April 2024.

**Table 2:** Recruitment and Retention.

| **Site** | **Number screened** | **Participants who were recruited** | **Participants randomised to control arm** | **Participants randomised to MATILDA arm** | **Participants attending community group at 6-months** | **Participants attending community group at 12-months** |
| --- | --- | --- | --- | --- | --- | --- |
| **N Ireland** | 55 (56%) | 33 (58%) | 17 (59%) | 16 (55%) | 13 (64%) | 13 (72%) |
| **London** | 40 (44%) | 24 (42%) | 12 (41%) | 12 (45%) | 6 (36%) | 5 (28%) |
|  | **95** | **57 (60%)** | **29** | **28 (\*25)** | **19 (76%)** | **18 (94.7%)** |

As shown in Table 2, of the 57 participants, 29 were randomised to the control arm and 28 were randomised to the MATILDA intervention arm. Of the 28 adults with disabilities randomised to MATILDA, two participants unexpectedly died and one fully withdrew. Our intervention arm therefore reduced from 28 to 25 adults with an intellectual disability across both sites.

### Matching participants to community groups / mentor(s)

We were able to match all 25 participants to a community group (100%) (such as Art & Crafts groups, Walking groups, Church Bowl groups, Evening Social Club, Lego club, Men Shed’s, Cuppa & Chat groups, Coffee Morning groups, or a Singing group). These groups were offered from biweekly, weekly, fortnightly to monthly depending on the time of the year and weather.

We were able to match 23 participants with at least one mentor (92%). Most participants were matched to one mentor, although some were matched to 2 mentors in their local community group. One man in Northern Ireland was matched to a local Men’s Shed, and one man in London was matched to a community group, although the community groups did not want to identify a specific mentor. However, all the members of the two community groups offered their support to the person with the intellectual disability on a weekly basis to embed them into the group. Two of the adults were attending their community groups weekly and continued to attend at the 12-month follow-up period.

### The matching process

> *“I think certainly the volunteer coordinator’s input was very good as in it wasn’t a case of bringing X along one week and going, there you go, leaving them alone…they fully supported them.”* (Disability Staff Member)

The matching process took longer than anticipated in many cases: we had originally planned 2-4 weeks; but in fact, this took up to 12-weeks. Part of this increased timeframe can be attributed to the time taken to identify community organisations, ensure they were suitable environments, and to explain the MATILDA intervention to group members (potential mentors) in sufficient detail. The VC’s also physically supported the person with the disability to attend the community group alongside the mentor(s) for several weeks, until the participant and mentor(s) became more comfortable with each other: the length of time depended upon each dyad’s circumstances (range 3-12 weeks). When confidence and trust were developed by both parties, the VC’s gradually withdrew their support over a 6-month period: again, this depended upon each person (range 5-26 weeks). For most adults with disability, they asked about the VC when the researcher was collecting the follow-up data at 6-and 12-months, a genuine testimony reflecting the trusting relationship that developed.

### Recruiting community groups

> *“Volunteer coordinator approached us to see if X could possibly attend a community group that I belong to with a view to sort of getting him integrated into the club and becoming part of it. The MATILDA Intervention was a bridge to make that introduction and hopefully then to be a long-term thing. So, not just part of the intervention for six-months. And that’s how it’s worked out for us. X is coming along now under his own steam and is an active part of the club.”* (Mentor)

Recruitment and reception of the community groups to the MATILDA intervention demonstrated notable variation across the two sites (see Table 3). In Northern Ireland, 30 older persons’ community groups were approached and most groups agreed to participate. One group declined due to unwillingness to complete the mentor documentation required, although the participant matched to this group continued to attend independently of the study. A second organisation declined due to instability of funding and inability to take on additional perceived work.

**Table 3:** Numbers of older persons community groups contacted.

| **Site** | **Total contacted** | **Agreed to take part** | **Declined** | **Other** |
| --- | --- | --- | --- | --- |
| **London** | 82 | 18 | 13 | 51 |
| **N. Ireland** | 30 | 28 | 2 | - |

The London site approached 82 community groups, 51 groups were ineligible for inclusion, due to post COVID-pandemic merging and dissolution of groups or a lack of long-term activities running (see Table 3). Among those 31 groups deemed eligible, thirteen groups declined to take part. Common barriers included the belief that involvement in the MATILDA intervention would be an additional burden on their group members, some community groups were understaffed or already at capacity for new members. Furthermore, there was noted apprehension regarding possible disruption to the existing group dynamics, with some community groups overestimating the risk associated with people with intellectual disability. Additional assurances were required to challenge biases held by group members, as well as clarification regarding practical concerns such as organisational liability and insurance coverage.

### Acceptability of the MATILDA Intervention

> *“I love mixing with people and they like me; I like to join the Bingo. The other members say hello to me, nice to see you. You know.”* (Older adult with an intellectual disability)

Findings from the focus groups indicated that the adults with disability clearly accepted the novelty of engaging with their local community group via the support of their mentor(s) where they both shared common interests. MATILDA was different to other community groups as no other people with an intellectual disability attended.

Further qualitative analyses of the focus groups with the adults with disability and their mentors revealed that this was due to: a) the structural characteristics of the community groups (i.e. sharing similar interests, close to home, appropriate in duration, small group size); b) the careful matching of the person with disabilities to a mentor(s) (i.e. age, gender, interests / hobbies); and c) the support of the VCs at both sites (i.e. a strong ‘implementation champion’ within the local community, providing ‘check-in points’ and offering continuity).

### Fidelity checks

> *“I thought the VC built a really good rapport with X. I really did. I thought it was lovely…It’s like you felt confident that she had thought of everything, you know what I mean?”* (Family member)

> *“I suppose when the VC was there, it was a bit of a safety blanket. Having someone there constantly beside her, I think that allowed her to be a bit more comfortable and settle in well.”* (Disability Staff Member)

Both VC’s in Northen Ireland and London continually contacted the adults with disability and their mentor by phone, text and ad-hoc visits to the person’s community group regularly over the six-months of the intervention. This also included contacting the family and/or paid carers and community group managers, to ensure the person with disability was regularly attending, mentor(s) were still facilitating the person to embed within the local group, and to identify and resolve any concerns.

These implementation strategies were deemed to be very successful by all as evidenced within the fidelity checks and focus groups with adults with intellectual disability, mentors, family and/or paid carers and managers.

### Adherence

Of the 25 older adults with an intellectual disability matched to a local community-based group, attendance records showed that 18 participants (72%) were regularly attending their group at 6-months. Moreover, 17 out of these 18 older adults were attending their community group (94%) regularly at the 12-months follow-up. A clear statement that the MATILDA intervention was positively working.

We had matched four older adults with an intellectual disability to a local community group, trained the mentors, and these participants were already attending these groups for several weeks. However, these four participants requested a rematch to a new community group as they described the current group / mentor(s) as being *‘too old / boring / not very posh’,* or experienced scheduling conflicts with existing activities such as day center / college. Three of these older adults with disability were re-matched successfully with an alternative local community group and mentor(s); however, one participant chose not to be re-matched due to ongoing physical health difficulties that they were experiencing. Another participant transitioned to an alternative local community singing group specific for people with dementia after receiving a dementia diagnosis.

### Reach

> *“I made a planter…growing lettuce and carrots and spuds and stuff like that”*. (Older adult with an intellectual disability)

> *“…we go on the trips and all with them (mentors), like garden centre or for lunch and we get away in the summertime, different places we go with them.”* (Older adult with an intellectual disability)

Data collected from the VC’s attendance records, alongside focus groups with the adults disability and their mentors, indicated that 12 of the 17 participants (71.5%) engaged in other social activities in addition to engaging with their local community group. Additional activities included involvement in local events (e.g. visits to garden centres, luncheon clubs, bingo, theatre performances, and Christmas pantomimes), and the development of new social connections with individuals outside of their residential setting without disabilities. The adults with intellectual disability also reported acquiring new skills, such as knitting, playing indoor bowls, engaging in creative and craft-based activities, and basic woodwork. This clearly demonstrated the wider community reach and engagement that MATILDA has offered these adults with an intellectual disability. A potential implementation strategy to ensure the older adults with disability remained in their community group after the VC’s withdrew at 6-months was the development of ‘succession plans’ with all stakeholders. These ‘succession plans’ were arranged before the end of the 6-month period and was key to the longevity of the intervention.

### Barriers to engaging with local community groups

> *“It cost me £20 for the taxi to go to the group, from my house down to X and then from X back up to my house”*. (Older adult with an intellectual disability)

> *“Unfortunately, person in the same group home] he cannot stay home as well without the staff in the building…”* (Disability Staff Member)

Using data collated from the focus groups, attendance records, and engagement with the VCs, there were several implementation barriers that prevented some of the adults with disability from regularly attending their local community group or stopped them attending altogether. These ranged from deteriorating in personal health, low mood / motivation, holidays, transport, and the group clashing with other appointments / activities. Another core reason for three older adults with intellectual disability stopping attending their local community group, was that the disability staff within their supported living scheme could no longer provide support and/or transport to attend their community group.

### Lost to follow-up

All 57 adults with intellectual disability completed the baseline questionnaires. By the time of the 6-months follow-up, sadly three participants had died (two had been randomised to the intervention arm but had not commenced their community group, and one person who died was from the control arm). Another person withdrew completely due to multiple family bereavements, and another person was uncontactable at this data collection point. That meant 52 people had their 6-month data collected (91.2%).

At the 12-months follow-up, one person had developed sudden onset dementia, and we were unable to collect the data at this time point. Two adults with intellectual disability were uncontactable and another person refused to engage in the 12-month data collection: a total of five participants. That meant, of 52 people who had their 6-month data collected (91.2%), we lost another three participants at this 12-month data collection point (94.2%). All the adults with intellectual disability indicated that the data collection process (i.e. time to needed to complete questionnaires, supports (researcher and/or carers) available to complete questionnaires), and the easy read versions of the questionnaires themselves, were all acceptable and helpful.

### Mentor’s completion of questionnaires

With regards the mentors, we only were able to get two mentors to complete their baseline questionnaires across both sites. We discussed the challenges with this low response rate with the two VCs and with the mentors themselves during community group visits. Potential reasons for non-completion included age of mentors (over 60 years of age, poor levels of literacy, not understanding why these questionnaires were needed and additional burden on mentors). Therefore, to minimise the burden on mentors and as a means of encouraging completion at the 6- and 12-month follow-up data collection points, we decided to reduce the number of items on each questionnaire based upon the factor loadings and the questions most relevant to the mentor group. We also attempted to give the questionnaires out at the focus groups at the 6-month follow-up.

The WHO QoL questionnaire examined four domains. We kept all four domains and selected items based upon the highest factor loadings and/or item most relevant to the aims of the intervention. We reduced the items to four items per sub-scale, in total 16 items. Factor loadings were examined using the work of (44) with the most relevant items at face validity extracted.

The Attitudes to People with Learning Disability questionnaire examined four domains, we removed one sub-scale related to the causation of intellectual disability as this was not relevant to the MATILDA intervention. We observed the factor loadings of the other three domains and reduced the number of items to 8, keeping those items with highest factor loadings and/or most relevant to aims of the intervention. This was based on the work of Bollen who wrote that if questions are removed from a one factor model, the factor is still a factor (46). Despite our efforts, only 8 mentors completed their 6-month follow-up questionnaires, and only 4 mentors completed their 12-month follow-up questionnaires.

### Clinical Outcome Data

Table 4 shows that the Glasgow Anxiety Scale scores for the participants in the MATILDA arm decreased from a mean of 20.7 at baseline to a mean of 18.1 at 6-months and a mean of 16 at 12-months. This is compared to the participants in the control arm who displayed a slight decrease across the three time-points (baseline mean (19.6), 6-month mean (17.1) and 12-month mean (18.2)).

**Table 4:**
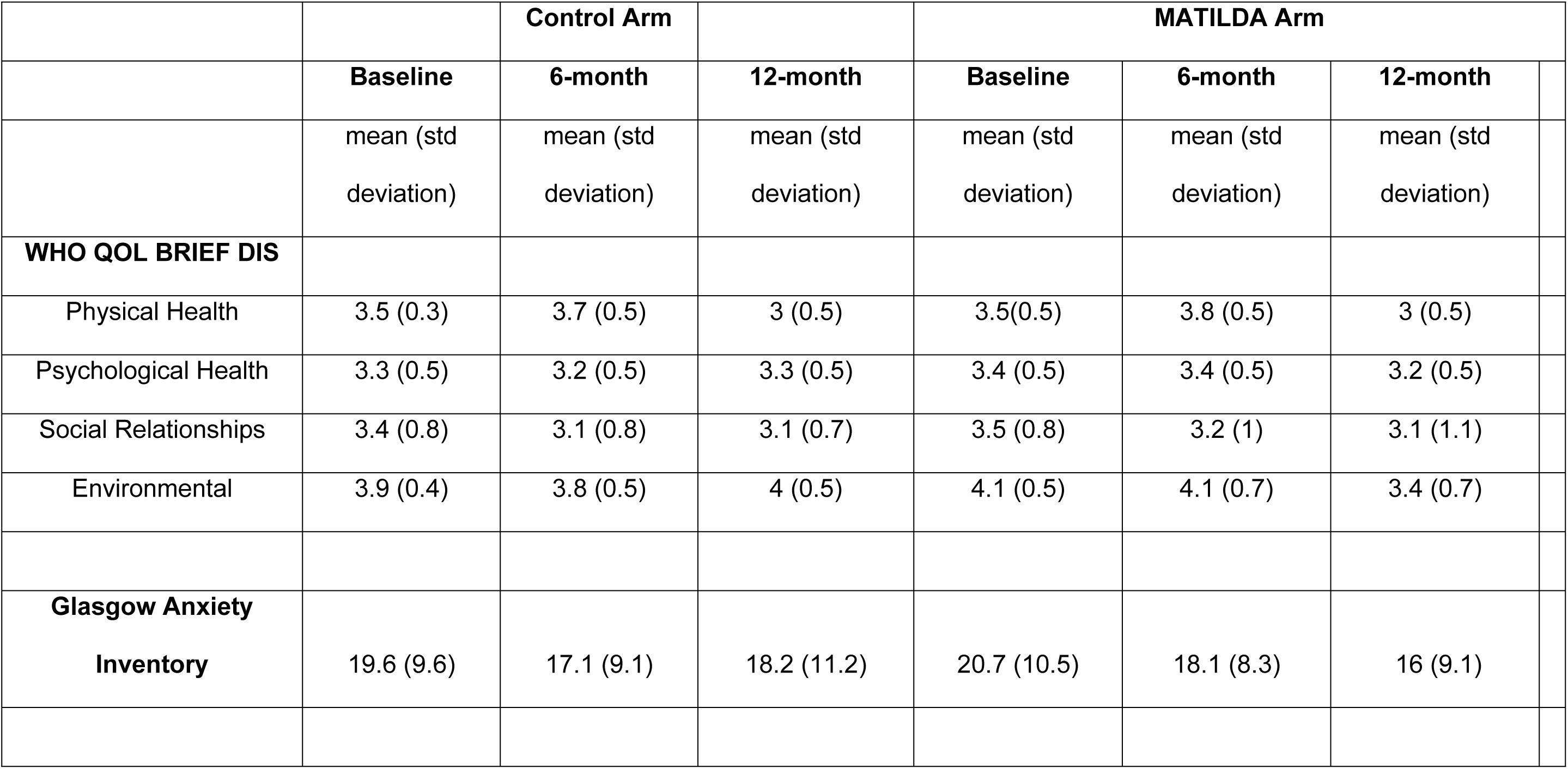
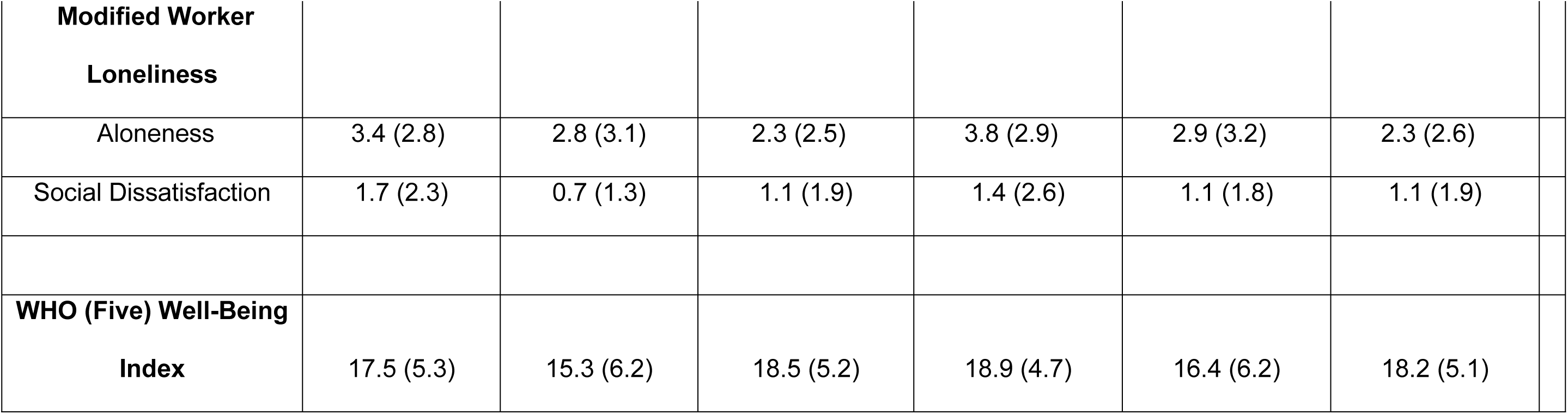
Mean and standard deviations of the secondary outcome measures for the participants in the MATILDA and Control Arms.

It can be observed in Table 4, that there were minimal differences in the participants scores across the four domains of the WHO-QOL DIS (physical health, psychological health, social relationships, and environment QOL) across the three-points (baseline, 6- and 12-months) for those in the MATILDA and control arms.

Table 4 shows that the Modified Worker Loneliness Aloneness sub-scale scores for the participants in the MATILDA arm decreased from a mean of 3.2 at baseline to a mean of 2.9 at 6-months and a mean of 2.3 at 12-months. This is compared to the participants in the control arm who also displayed a decrease across the three time-points (baseline mean (3.4), 6-month mean (2.8) and 12-month mean (2.3)). With regards the Modified Worker Loneliness Social Dissatisfaction sub-scale there was minimal differences in scores across the three time points for participants in both arms.

Likewise, there were minimal differences in the participants scores across both the Matila and control arms on the WHO (Five) Well-Being Index and EQ-5DY across the three time-points.

### Health Economics

The data indicates that it is possible to collect health-related QOL data and data on aspects of service use. It was clearly feasible to collect this data across both sites, and up to and including the final 12-month time point. That a range of values are recorded using HR-QOL instruments suggests: first, that there is real engagement with instruments rather than token engagement as might be indicated by clustering in responses or a high level of non-response; second, the instruments can differentiate between individuals in terms of HR-QOL, that is they appear not to be subject to ceiling or floor effects and; third, the low number of missing values at this third time point remains low, suggesting attrition may not be a major factor. These findings are mirrored across both sites.

### Adverse events

There were three sudden deaths, but these events were not related to the intervention.

## Discussion

The study reports on the outcomes of the MATILDA feasibility RCT, a novel community-based intervention targeting older adults with an intellectual disability to engage in a local community group with support from trained mentors. We used the RE-AIM framework (32) to guide the process evaluation and explore the feasibility of the clinical outcomes.

Our results revealed that we can clearly identify, recruit, consent, randomise, and match adults with an intellectual disability to a community group with a mentor(s). Implementation of MATILDA was evaluated by determining whether the intervention was delivered as intended. The percentage of community group activities attended was deemed to be moderate to high for most of the adults with intellectual disability. We found 18 of the 25 adults with intellectual disability (72%) where still attending their community groups at 6-months, and 17 of the 18 participants still attending at the 12-months (94%). Furthermore, many of the participants were found to widen their social connections by attending additional community activities and events.

These results clearly illustrate the reach and sustainability of the MATILDA intervention.

Our results echo the work undertaken in the ‘Transition to Retirement Intervention’ in Syndey, Australia who reported that 25 of 29 (86%) older adults with an intellectual disability in the intervention were still attending their group at 6-months (30). There was no formal 12-month follow-up, however Stancliffe et al. noted that *“most participants continued to attend their group beyond cessation of data collection”* (p. 711).

Conversely, our results clearly differed from the earlier Ali et al (2021) feasibility RCT of a befriending scheme that found it was difficult to identify, consent and match community volunteers to adults with intellectual disability (47).

Through the focus groups with the mentors and community group managers, we found that MATILDA was deemed acceptable. This was strongly associated with the following mechanisms: a) the structural characteristics of the community groups b) the careful matching of the person with disabilities to a community group and mentors and c) the support of the two VCs who acted as strong *‘implementation champions’* at each site. In addition, using similar group-based arts / hobbies / activities that align with both the adult with intellectual disability and the mentors / wider groups’ interests offers valued social identity and a shared focus. Both the person with the disability and mentors / wider group members are accepted for their valuable contribution rather than their disability creating a collective sense of purpose and belonging (48).

This success can be further explained by the VC’s building strong trusting relationships and clear open channels of communication with 1) the community group managers and mentors and 2) the adults with intellectual disability and their family/paid carers throughout the matching process and during the 6-months of the intervention (by phone, text and ad-hoc visits). Adoption was demonstrated in how the mentors, community group leaders and wider community group members, embraced their supportive roles offering practical and emotional support that enabled the older adults with intellectual disability to participate in the local groups expanding friendships, social networks, building confidence, and reducing social isolation and depression (49, 50). This willingness to adopt supportive roles laid part of the foundation for effective implementation, shaping how MATILDA was successfully delivered and sustained in practice.

Patterns of adoption also have implications for maintenance, as the sustained engagement from mentors, community leaders and wider group members at 6-months is likely to have influenced the long-term embedding of the intervention within the community groups at 12-months.

These results are comparable to Wilson et al (2013), Bigby et al. (2014), and Stancliffe et al (2015) who identified that the success of the Australian ‘Transition to Retirement’ intervention was based upon the co-ordination with multiple stakeholders, the significant time spent by the VC’s negotiating access, matching, and supporting the adults with intellectual disability into community groups and matching to mentors (49, 51, 30). Likewise, the sustained facilitation of the two VCs also found at the two sites helped build familiarity, motivation, willingness, confidence and resolve anxieties to make inclusion work with the mentors in the community groups for the adults with intellectual disability.

The mentors offered enough active support, practical and emotional support, opportunities, leading the adults with intellectual disability to needing less support over time, increasing self-confidence, independence, less anxiety, and lasting connections and friendships. This subsequently led to stable participation, positive relationships, successful role integration, and long-term maintenance of engagement in the community groups. Amado et al. (2014) reported that such mechanisms provided by the VCs, mentors and wider group membership enhanced greater feelings of group cohesion, genuine belonging and self-worth for the adults with disability thereby making community-based groups more acceptable and inclusive (50).

This study has shown that the clinical outcome measures (quality of life, anxiety, loneliness) were acceptable to the adults with intellectual disability and we had minimal missing data across the three-time points. It is worth noting that these standardised questionnaires had been developed for adults with disability. However, the questionnaires were not acceptable to the mentors despite our efforts to shorten them.

Although not statistically powered, for those adults with intellectual disability in the MATILDA arm there was some improvement in their anxiety scores across the three-time points compared to the participants in the control arm. Therefore, MATILDA has the potential to improve well-being, anxiety and social connectedness for older adults with intellectual disability in a larger powered study. We also found that we could collect health-related QOL data and data on aspects of service use. These results clearly demonstrate that we met our progression criteria for a definitive RCT to examine the clinical and cost-effectiveness of the MATILDA intervention for older adults with an intellectual disability can now proceed.

Through the interviews and focus groups, and VC’s logs, we found out there were several implementation barriers that prevented some of the adults with intellectual disability from regularly attending their local community group or stopping attending altogether. One implementation barrier was that some disability staff could no longer support them attend and/or provide transport to support them to attend their community group. This was due to several related organisational and staffing issues: high staff turnover and low morale of staff, poor communication about the purpose of the MATILDA intervention, staffs’ commitment to support the participants, not being able to provide transport. Abbott and McConkey (2006) reported that paid support staff may adopt paternalistic, risk-averse approaches that inadvertently restrict social opportunities for adults with intellectual disability, thereby limiting their autonomy and reducing their engagement in community life (52).

Kelly et al (2025) reported similar findings in their recent internal pilot RCT of a diabetes education programme for adults with intellectual disability across the UK (53). They discovered that service providers and disability staff sometimes struggled to support participants’ engagement in the nine-week diabetes education intervention. This was due to a lack of “buy-in” from senior managers, insufficient staff capacity and a lack of understanding of the study’s purpose and methodology. Kelly and colleagues recommended identifying these barriers to embedding the intervention earlier and using implementation theories and frameworks like the Consolidated Framework for Implementation Research 2.0 (Damschroder et al. 2022) and/or the Normalisation Process Theory (May et al. 2007) (54, 55) to address such barriers. Kelly and colleagues suggested matching these barriers to appropriate implementation strategies (Expert Recommendations for Implementation Change (ERIC): Powell et al. 2015).

## Conclusion

This feasibility RCT and process evaluation, guided by the RE-AIM framework (Acceptability, Reach, Effectiveness, Adoption, Implementation and Maintenance), revealed that the MATILDA intervention is a feasible community-based approach to support older adults with an intellectual disability in participating in their local community group. These findings provide valuable information that allows us to move forward with a definitive randomised control trial involving a larger and more diverse population. Furthermore, these results indicate that the MATILDA intervention offers a novel community-based, mentor-supported intervention for promoting social inclusion, wellbeing and sustainability.

## Data Availability

The data underlying the results presented in the study are available from (include the name of the third party and contact information or URL).

---

**Fig 1:**
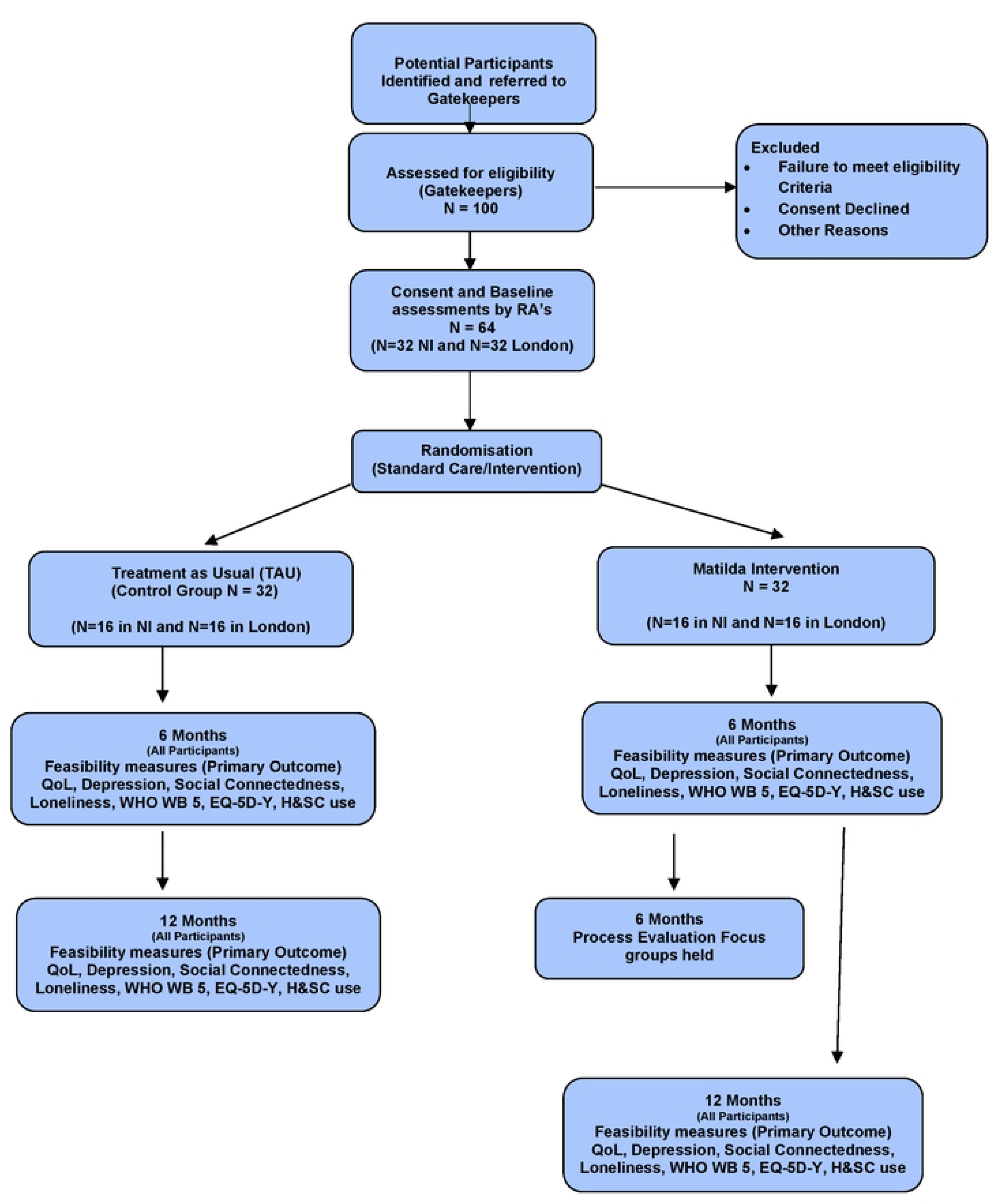
MALIDA Consort Diagram

